# Identification of risk and protective human leukocyte antigens in COVID-19 using genotyping and structural modeling

**DOI:** 10.1101/2021.05.04.21256636

**Authors:** Yiran Shen, David A. Ostrov, Santosh Rananaware, Piyush K Jain, Cuong Q. Nguyen

## Abstract

COVID-19 is caused by severe acute respiratory syndrome-coronavirus-2 (SARS-CoV-2). The severity of COVID-19 is highly variable and related to known (e.g., age, obesity, immune deficiency) and unknown risk factors. Since innate and adaptive immune responses are elicited in COVID-19 patients, we genotyped 94 Florida patients with confirmed COVID-19 and 89 healthy controls. We identified an HLA gene, HLA-DPA1, in which specific alleles were associated with the risk of SARS-CoV-2 positivity and COVID-19 disease. HLA-DPA1*01:03 was associated with reduced incidence of SARS-CoV-2 positivity, whereas HLA-DPA1*03:01 was associated with increased risk of SARS-CoV-2 positivity. These data suggest a model in which COVID-19 severity is influenced by immunodominant peptides derived from SARS-CoV-2 preferentially presented by specific HLA-DP molecules to either protective (for asymptomatic COVID-19) or pathogenic T cells (in severe COVID-19). Although this study is limited to comparing SARS-CoV-2 positive and negative subjects, these data suggest that HLA typing of COVID-19 patients stratified for disease severity may be informative for identifying biomarkers and disease mechanisms in high-risk individuals.

## Introduction

Classical human leukocyte antigen (HLA) genes (HLA-A, -B, -C, -DR, -DQ, -DP) exhibit a high degree of polymorphism and play critical roles in the immune response to viral infections. CD4^+^ T cells and CD8^+^ T cells respond to pathogens by recognizing different classes of HLA molecules (I or II, respectively) on the cell surface. Specific HLA genotypes have been associated with T-cell mediated immunity and viral clearance, while low-affinity peptide binding and antigen presentation may also make specific genotypes a risk factor for infectious disease. For example, HLA-A*02:01, -A*03:01, -B*08:01, -B*18:01, -B*37:01, -B*57:01, and -DRB1*09 alleles are involved in general protective CD8^+^ or CD4^+^ T cell-mediated immunity through presentation of conserved influenza peptides^1–4^. In contrast, HLA-A*11, -A*24, -A*68, -B*35, and -DRB1*10 alleles may be the associated risk factor for severe pandemic influenza A (H1N1) infection and another common circulating influenza strains^5–7^. Studies of SARS-CoV demonstrated that HLA-B*46:01, -B*07:03, -Cw*08:01, -DRB1*12,:02 and -DRB4*01 alleles were associated with disease susceptibility or severity in various populations^8–12^. Similarly, HLA-DRB1*11:01 and -DQB1*02:02 alleles were associated with the risk of Middle East Respiratory Syndrome (MERS). HLA-associated adaptive immune responses were less efficient against human immunodeficiency virus (HIV)-1 than respiratory viruses^13^, but HLA-B*27, -B*57, -B*58:01 and -B*81:01 alleles were found to present multiple p24 Gag-specific epitopes in different ethnicities and confer protective effects against HIV disease progression, whereas HLA-B*42:02 and -B*58:02 alleles were associated with susceptibility and rapid disease progression^14^.

Predictions of binding affinity between HLA class I and II molecules and peptides of SARS-CoV-2 proteins showed that specific alleles might be associated with disease morbidity and mortality. For example, consistent with a previous study of SARS-CoV, HLA-B*46:01 allele was predicted to be associated with SARS-CoV-2 susceptibility using silico analysis of viral peptide-MHC class I binding affinity to HLA-A, -B, and -C genotypes for all SARS-CoV-2 peptides ^15^ HLA-C*05 alleles were associated with risk of death from COVID-19^16^. Conversely, HLA-A*02:11, -A*02:22, -B*15:03, -DRB1*01:01, and -DRB1*10:01 alleles were predicted to be protective due to their enhanced ability to present SARS-CoV-2 peptides^13,15,17^. Using whole-genome sequencing, a recent study with 332 patients demonstrated that HLA-A*11:01, -B*51:01, and -C*14:02 alleles were correlated with disease severity^18^. These early studies suggest that HLA alleles are related to susceptibility and disease prognosis. Thus, identifying the association with SARS-CoV-2 positivity will help clarify the heterogeneity of responses to the disease, potentially guiding personalized treatments and developing epitope-based peptide vaccines against SARS-CoV-2. In this study, we sought to determine if specific HLA alleles were associated with susceptibility to SARS-CoV-2 infection/COVID-19 disease by comparing SARS-CoV-2 positive subjects in a Florida population with SARS-CoV-2 negative subjects from the same population.

## Materials and Methods

### Study population

Ninety-four confirmed COVID-19 samples were obtained from Boca Biolistics (Pompano Beach, FL) and CTSI Biorepository at the University of Florida (Gainesville, FL). The median age of the patients was 55.7□years (range: 3-94□years). Patients had positive test results for SARS-CoV-2 by RT-PCR from nasopharyngeal swabs or tracheal aspirates. Eighty-nine healthy individuals who had negative test results at the same sites and time were included as controls. The median age of the control group was 60 years (range: 0-101 years). In the SARS-CoV-2 positive cohort, there were 50 females, 43 males, and 1 undisclosed, in which there were 17 white, 21 black, 1 Asian, 1 Non-Hispanic, and 55 undisclosed. In control SARS-CoV-2 negative cohort, there were 47 females and 42 males, in which there were 44 white, 10 black, 1 Non-Hispanic, and 34 undisclosed enrolled. The study was approved by the Institutional Review Board of the University of Florida.

### HLA allele typing

Clinical specimens of nasopharyngeal swabs were collected in a viral transport medium. DNA was extracted from viral transport medium or directly from tracheal aspirates by Maxwell® RSC Blood DNA Kit per manufacturer’s instructions (Promega Corporation). RNase A was added to samples to remove potential viral RNA. Isolated genomic DNA was quantified by NanoDrop™ One/OneC Microvolume UV-Vis Spectrophotometer (Thermo Scientific). Genotyping was done using Axiom™ Human Genotyping SARS-CoV-2 Research Array as instructed by the manufacturer (Thermo Scientific). Genotyping data were assessed by Axiom™ Analysis Suite Software and filtered according to QC and HLA-related datasets. Hardy-Weinberg Equilibrium (HWE) was evaluated for all the SNPs evaluated, and all SNPs selected for HLA allele typing meet the criteria (p > 0.05). Automated high-resolution HLA typing was done by Axiom™ HLA Analysis Software, and certain HLA-A, -B, -C, -DPA1, -DPB1, -DQA1, -DQB1, -DRB1/3/4/5 alleles of each loci were assigned to samples.

### Peptide binding analysis

The Wuhan-Hu-1 sequence of the spike glycoprotein was used to predict peptides that bind HLA-DP molecules (https://www.ncbi.nlm.nih.gov/datasets/coronavirus/proteins/). The SMM-align (NetMHCII-1.1)(PMID:17608956) prediction method implemented in IEDB was used to generate the half-maximal inhibitory concentration (IC50) values that estimate dissociation constant (K_D_) in nanomolar values.

### Structural modeling of spike peptide HLA-DP interactions

The crystal structure of HLA-DP complexed with a peptide corresponding to RAS (PDB code 4P5K, 24995984) was used for modeling the SARS-CoV-2 spike peptide VVFLHVTYVPAQEKN (positions 1060-1074, corresponding to the S2 subunit). The HLA-DP α-chain in this structure corresponds to HLA-DPA1*01:03:01^19^. The RAS peptide NKFDTQLFHTITGGS was mutated to VVFLHVTYVPAQEKN using COOT^20^ with rotamers that represent a local energy minimum of torsional angles. The geometry of the resulting complex was regularized in PHENIX^21^. The amino acid sequence of HLA-DPA1*03:01:01 from IMGT^19^ was used as the basis for structural modeling with SWISS-MODELER, which generated a protein data bank format file based on a crystal structure of HLA-DP α-chain 98.9 % identical to HLA-DPA1*03:01:01, PDB 4P5M. PyMOL (https://pymol.org/2/) was used to generate molecular graphic images.

### Statistical analysis

HLA class I and class II allele frequencies were estimated by direct counting based on results from allele typing, calculated as the ratio of the number of times different alleles appeared in the sample to the total number of alleles.. Odds ratios (ORs, 95% confidence interval [CI]) and p-values were calculated using R 4.0.3 (R Core Team, 2018), the exact2×2 (v1.6.5; Fay MP, 2010) package or Prism 9.0 software (GraphPad Software, La Jolla California USA). 0.5 was added to all the cells to prevent computation errors when calculating the odds ratio or standard error. When zero values caused computational problems with the odds ratio or its standard^22^. Since small size samples were used in this analysis, the risk of introducing a bias in estimating the probability (p-value) and wrongly accepting association (type I error) was corrected using the method of Benjamini-Hochberg. In this method, the p-value is multiplied by the number of alleles input at each locus, thereby giving a more powerful corrected p-value (pc-value) that may be interpreted with confidence.

## Results

### HLA typing of COVID-19 patients

We genotyped 94 SARS-CoV-2 positive patients and 89 healthy control populations matched as control. We identified multiple alleles present for HLA-A, -B, -C, -DPA1, -DPB1, -DQA1, -DQB1, -DRB1 loci (30, 46, 20, 5, 20, 7, 16, and 32, respectively (**Table S1**)). HLA-DRB/3/4/5 were not analyzed due to high rates of ambiguous imputation (**Table S2**). The allele distributions of HLA-A, -C, -B, -DRB1, -DQB1, and -DPB1 loci were compared between COVID-19 patients and control individuals. The frequencies and odds ratios (OR) of HLA alleles with significant associations with incidence of SARS-CoV-2 positivity are shown in **Table 1**. Potentially significant associations between HLA-DPA1 alleles and SARS-CoV-2 positivity were identified. DPA1*03:01 was associated with an increased risk of SARS-CoV-2 positivity (OR 9.3, CI 1.3 - 200.4, p=0.01). DPA1*03:01 is a rare allele (frequency < 1% in controls), but present in 6% of SARS-CoV-2 positive individuals (Pc value 0.06). In contrast, DPA1*01:03 was associated with reduced incidence of SARS-CoV-2 positivity (OR 0.6, CI 0.4 - 0.9, p=0.02). DPA1*01:03 is a common allele (frequency 72% in controls), but less frequent in individuals infected with SARS-CoV-2 (60 %, Pc value 0.06).

**Table 1.** Significant alleles associate with either protective (OR <1) or risk (OR>1) factor.

| Allele | COVID-19 |  | Control group |  | Odds ratio<br>(95% CI) | P-value | Pc-value |
| --- | --- | --- | --- | --- | --- | --- | --- |
|  | N | Frequency (%) | N | Frequency (%) |  |  |  |
| **DPA1\*03:01** | 10 | 5.9524 | 1 | 0.6757 | 9.2568(1.3264 - 200.3514) | 0.0120 | 0.06 |
| **A\*74:01** | 9 | 5.4878 | 1 | 0.6757 | 8.4924(1.2738 - 185.6196) | 0.0211 | 0.579 |
| **A\*30:01** | 8 | 4.8780 | 1 | 0.6757 | 7.5017(1.1526 - 165.9402) | 0.0386 | 0.579 |
| **DPB1\*01:01** | 23 | 15.5405 | 8 | 5.7971 | 2.9792(1.2446 - 7.258) | 0.0123 | 0.214 |
| **DQA1\*05:01** | 40 | 26.3158 | 22 | 15.9420 | 1.8791(1.0435 - 3.5288) | 0.0326 | 0.1141 |
| **DPA1\*01:03** | 10<br>1 | 60.1190 | 107 | 72.2973 | 0.5786(0.3551 - 0.9401) | 0.0244 | 0.06 |
| **DQA1\*02:01** | 14 | 9.2105 | 26 | 18.8406 | 0.4383(0.2122 - 0.8942) | 0.0257 | 0.1141 |
| **DPB1\*11:01** | 2 | 1.3514 | 9 | 6.5217 | 0.1974(0.0301 - 0.9301) | 0.0301 | 0.396 |

### Race comparison of risk and protective HLA

It is well established that minority groups of different races and ethnic groups in the US are disproportionately affected by COVID-19. Minorities endured a higher risk for infection, hospitalization, and death^23,24^. Therefore, in this study, we sought to determine if there was an association between the identified HLA and SARS-CoV-2 positivity in different races in the Floridian population. The data indicated that HLA-A*02:01 was associated with reduced incidence of SARS-CoV-2 positivity (OR 0.1, CI 0.003-0.6, p=0.02) in the black population (**Table S3**). Black individuals carrying the common HLA-A*02:01 allele were ten times more likely to test negative for SARS-CoV-2 than black individuals lacking HLA-A*02:01. Similar analyses were performed for the white Floridan population. As presented in **Table S4**, there was no significant allelic association in this population. The data indicate that HLA-A*02:01 allele might be protective for the black minority group.

### Structural modeling of HLAs binding to SARS-CoV-2 spike protein

Since different allelic forms of HLA molecules exhibit binding preferences for distinct sets of antigenic peptides, specific HLA allotypes may be less likely to present certain peptides derived from SARS-CoV-2 to T cells, resulting in a weaker antiviral response (as suggested by the association of HLA-B*46:01 in SARS patients^8^). In contrast, other HLA molecules may bind with higher affinities to immunodominant SARS-CoV-2 derived peptides, resulting in more robust antiviral responses. Since the dominant immunogenic T cell epitopes derived from SARS-CoV-2 were recently defined from all proteins, including in the critical spike protein^25^, we sought to identify peptides that bind preferentially to HLA molecules associated with risk or protection. We identified a peptide corresponding to the SARS-CoV-2 spike protein (15 amino acids, VVFLHVTYVPAQEKN) predicted to bind HLA-DP molecules related to risk with different affinities **(Figure 1)**. VVFLHVTYVPAQEKN was predicted to bind an HLA-DP molecule associated with reduced incidence of SARS-CoV-2 positivity (i.e., lower levels of infection) with high affinity (estimated Kd 298 nM for HLA-DPA1*01:03 α-chain/HLA-DPB1*02:01 β-chain, IEDB). In contrast, the same peptide was predicted to bind an HLA-DP molecule associated with increased risk of SARS-CoV-2 positivity with a low binding affinity (estimated Kd 1.7 μM, HLA-DPA1*03:01:01 α-chain/HLA-DPB1*04:02 β-chain).

**Figure 1.**
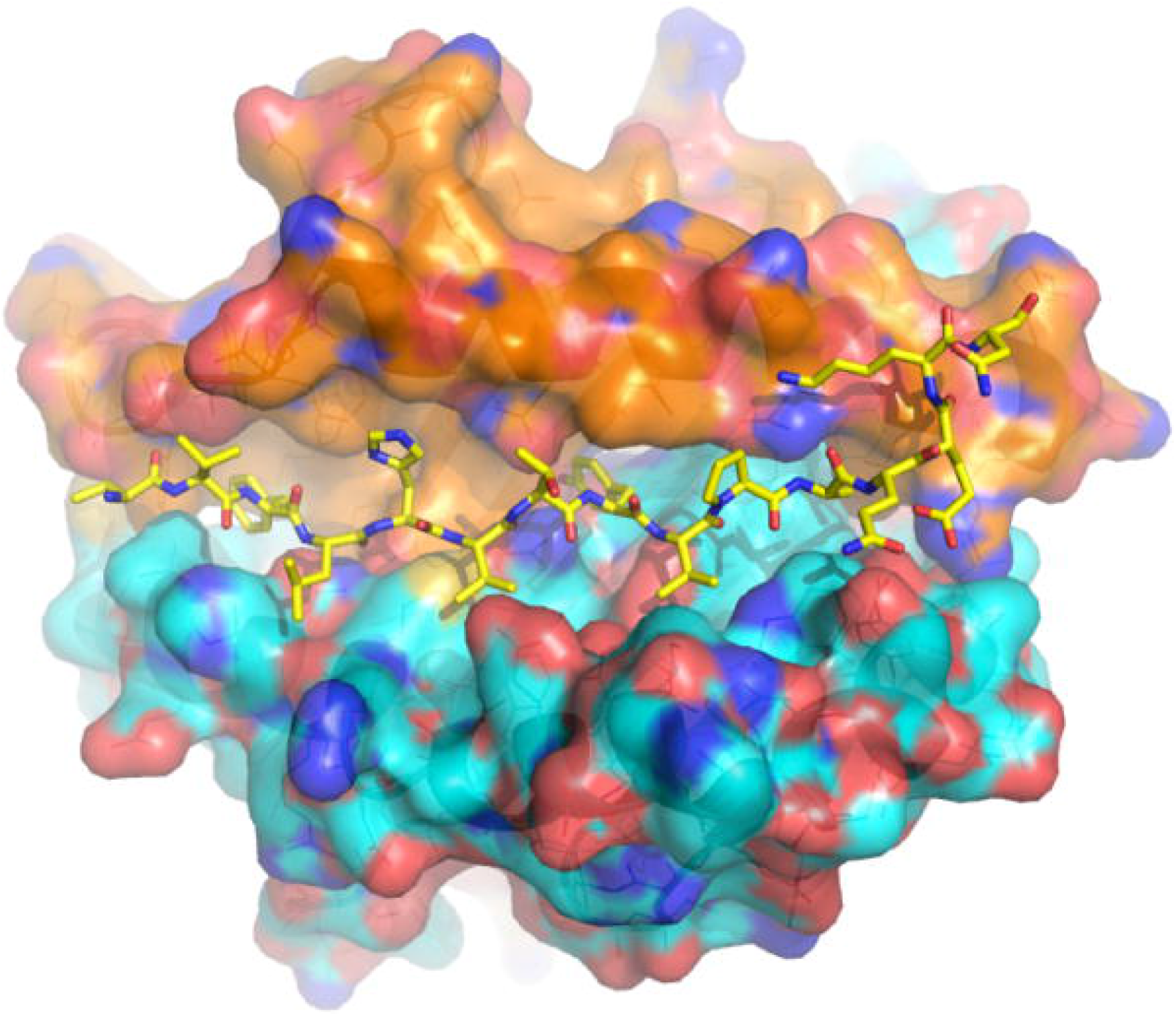
Model of an HLA-DP molecule associated with reduced incidence of SARS-CoV-2 positivity. SARS-CoV-2 spike peptide VVFLHVTYVPAQEKN, is shown as sticks modeled on the crystal structure of HLA-DP (PDB code 4P5K, DPA1*01:03 α-chain), yellow for carbon, red for oxygen, blue for nitrogen. The molecular surface of the HLA-DP α-chain is shown in orange for carbon, red for oxygen, blue for nitrogen. The molecular surface of the HLA-DP β-chain is shown in cyan for carbon, red for oxygen, blue for nitrogen.

Structural modeling suggests that SARS-CoV-2 spike peptide VVFLHVTYVPAQEKN binds DPA1*01:03 with higher affinity than DPA1*03:01 due to a polymorphism at position 42 located on the α1 helix of the DP α-chain, oriented toward the peptide at the center of the antigen binding cleft. Alanine at position 42 of DPA1*01:03 α-chain is predicted to anchor peptide binding by intermolecular contact with tyrosine in the peptide core (VVFLHVT**Y**VPAQEKN)(**Figure 2**, upper panel). The bulky methionine at position 42 of DPA1*03:01 α-chain is predicted to impose steric clash with the peptide core (**Figure 2**, lower panel), preventing formation of a high-affinity immunogenic epitope. These data suggest a model in which the course and severity of COVID-19 disease are influenced by the presentation of immunogenic peptides by specific HLA molecules present in a subset of individuals, **Figure 3**.

**Figure 2.**
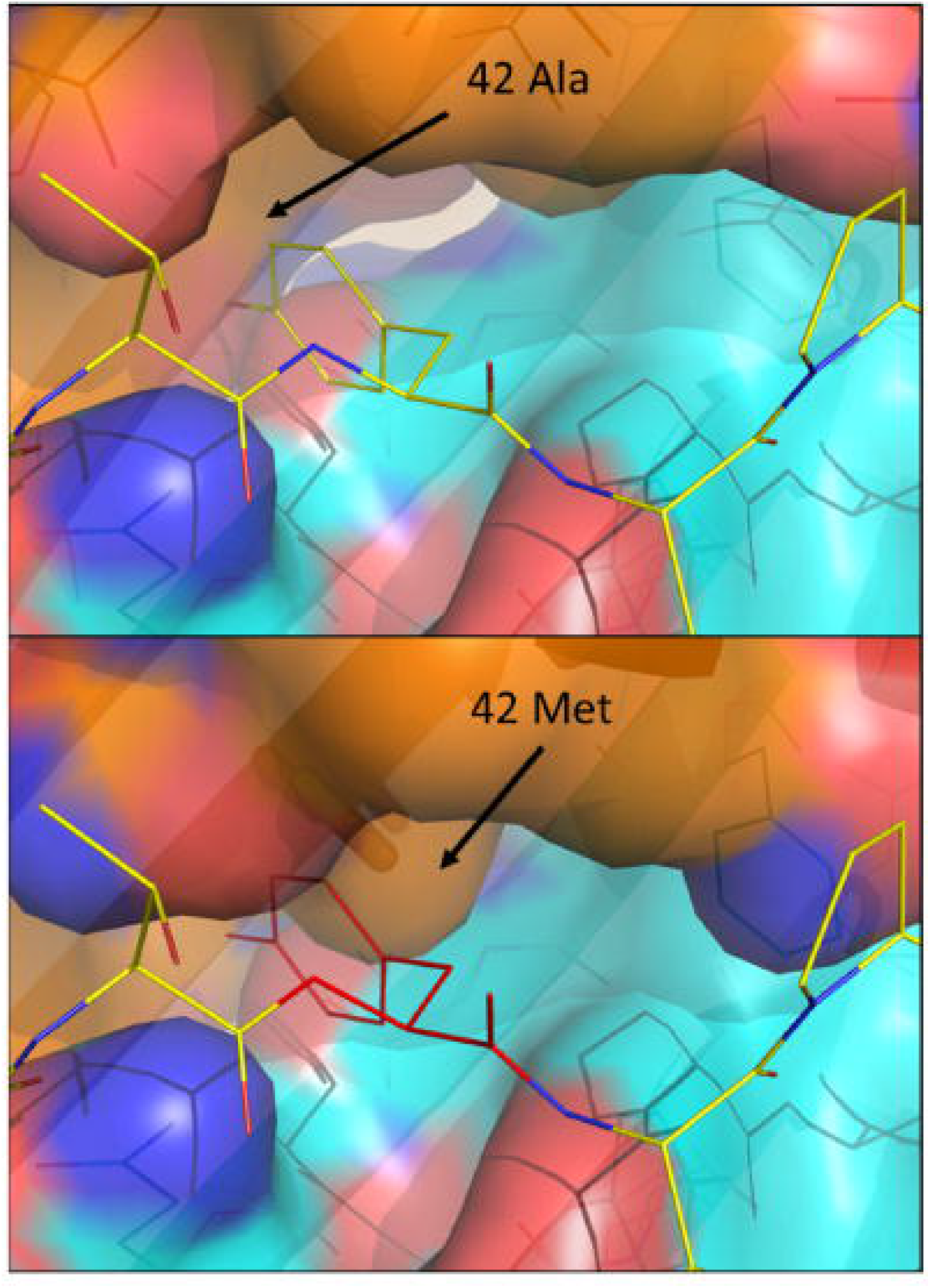
Polymorphism in the antigen binding cleft of HLA-DP has the potential to inhibit binding to peptides derived from SARS-CoV-2. Upper panel, DPA1*01:03, associated with reduced incidence of SARS-CoV-2 positivity forms an α-chain predicted to bind SARS-CoV-2 spike peptide VVFLHVT**Y**VPAQEKN with high affinity. The molecular surface of the HLA-DP α-chain is shown in orange for carbon, red for oxygen, blue for nitrogen. The molecular surface of the HLA-DP β-chain is shown in cyan for carbon, red for oxygen, blue for nitrogen. The tyrosine residue at the central peptide position forms intermolecular contact with alanine at position 42 of DPA1*01:03. Lower panel, DPA1*03:01, associated with increased incidence of SARS-CoV-2 positivity, forms an α-chain predicted to bind SARS-CoV-2 spike peptide VVFLHVT**Y**VPAQEKN with low affinity because of the steric clash between the tyrosine residue at the central peptide position (shown as red lines) and methionine at position 42 of DPA1*03:01.

**Figure 3.**
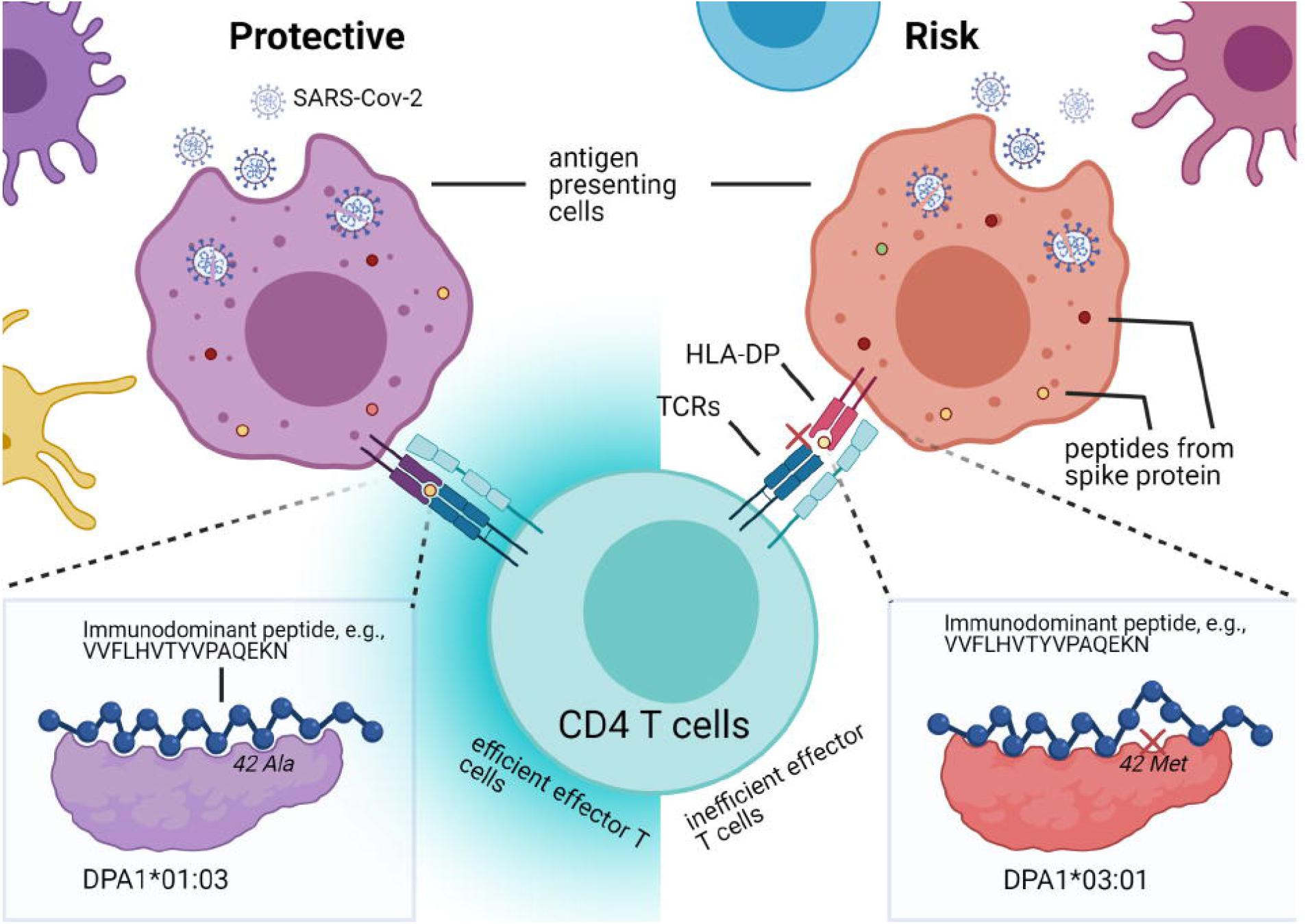
HLA-DP allele-specific viral clearance model. HLA-DP molecules with certain α-chains may bind immunodominant epitopes (color dots within cells) derived from antigen presenting cells with high affinity, whereas others may bind with low affinity, affects whether CD4 T cells can be effectively activated (left, efficiently activated; right, inefficiently activated) to promote downstream responses for viral clearance. Polymorphism at position 42, located on the α1 helix of the DP α-chain, leads to a difference in the binding affinity with peptides derived from spike protein (e.g., VVFLHVT**Y**VPAQEKN), alanine of HLA-DPA1*01:03 α-chain had intermolecular contact with tyrosine in the peptide core (lower left corner, the peptide is shown by ball-and-stick), and methionine of -DPA1*03:01 α-chain prevented high affinity binding with tyrosine due to steric clash (lower right corner, the peptide is shown by ball-and-stick, crossed at low binding affinity position). | Created with BioRender.com

## Discussion

In late December 2019, COVID-19 began spreading in Wuhan, Hubei Province, China, caused by a suspected zoonotic source of SARS-CoV-2^26,27^. While coronaviruses are relatively common, mutations can cause severe symptoms in humans; COVID-19 is the third noted case in which this has happened following SARS^28,29^ and MERS^30^. Globally, as of March 22^th^, 2021, there were more than 123 million confirmed cases and 2.71 million deaths worldwide; in the US there were 30 million confirmed cases with over 540,000 deaths^31^. Age and pre-existing medical conditions such as hypertension, obesity, chronic lung disease, diabetes mellitus, and cardiovascular disease are associated with disease severity and hospitalization rates of COVID-19 patients. In the COVID-19–Associated Hospitalization Surveillance Network (COVID-NET), which represents approximately 10% of the U.S. population with an equal frequency of males and females,, 54% of COVID-19-associated hospitalizations occurred in males and 46% occurred in females^32^. These data suggest that males may be disproportionately affected by COVID-19 compared with females. A meta-analysis of 15 independent studies documenting patient gender-specific outcomes found that men were more likely to develop severe COVID-19 infection than women (Odds Ratio, 1.31; 95% CI, 1.07-1.60)^33^. Other studies have indicated that black and Hispanic populations might be disproportionately affected by COVID-19^32^. The most interesting development in COVID-19 is that there are cases of children worldwide with COVID-19 exhibiting a Kawasaki-like disease^34–36^. Kawasaki disease has one of the strongest HLA associations, and there are significant differences in the distribution of HLA alleles among ethnicity^37^ This suggests that children with susceptible HLAs may develop Kawasaki-like disease with COVID-19.

There is a broad range of immunological responses to SARS-CoV-2, rendering individuals on a spectrum from asymptomatic to severely symptomatic for reasons that are not understood but likely result from genetic and environmental factors. To demonstrate whether HLA molecules are associated with COVID-19 infection and to further explore whether HLAs could serve as biomarkers for susceptibility or protection against SARS-CoV-2, we examined the HLA types of infected patients and compared them to samples collected from healthy individuals during the same period. The study identified that HLA-DPA1*01:03 was highly prevalent in healthy individuals but less in SARS-CoV-2 positive patients, and HLA-DPA1*03:01 was significantly associated in patients with SARS-CoV-2 infection. Using structural modeling with a potential T cell epitope derived from the spike protein, we predicted that HLA-DPA1*01:03 may be protective because of high binding affinity between SARS-CoV-2 peptides and HLA-DP. In our model, a polymorphic position in the center of the antigen binding cleft at position 41 in HLA-DPA1*03:01 was prevents high affinity peptide binding to HLA-DP, thus preventing specific T cell responses to SARS-CoV-2 peptides, consistent with risk associated with SARS-CoV-2 positivity.

HLA molecules present antigens by binding to endogenous antigenic peptides (class I) or exogenous antigenic peptides (class II) and express them as peptide-MHC complexes on the surface of antigen-presenting cells. During viral infection, cytotoxic T lymphocytes (CTL) kill viral-infected cells by recognizing HLA class I-peptide complexes at the cell surface, and CD4^+^ T cells recognize viral antigens presented by HLA class II molecules and activate antigen presenting cells to trigger an adaptive immune response against invading pathogens. Therefore, a spectrum of immune response is dictated by the flavor of the HLAs. Recent studies from different countries have identified multiple COVID-19 morbidity-related alleles; for example, Wang et al. identified HLA-B*15:27 alleles from a Chinese population^38^, Novelli et al. identified HLA-B*27:07, -DRB1*15:01 and -DQB1*06:02 alleles from an Italian population ^39^, Yung et al. identified serotype B22(HLA-B*54:01, B*56:01 and B*56:04 alleles) from Hongkong Chinese population^40^. To further support the concept that low affinity of viral peptides binding to HLA can predict susceptibility, Amoroso et al. have shown that HLA-DRB1*08 was correlated to mortality (6.9% in living versus 17.5% in deceased), and peptide binding prediction analyses demonstrated that these alleles were not able to bind SARS-CoV-2 peptides with high affinity. A similar finding was supported by a study with Sardian population in which HLA-DRB1*08:01 allele was found only in the hospitalized patients^41,42^. Structural modeling of SARS-CoV-2 spike peptides and HLA-DP interaction in our study demonstrated that a polymorphism at position 42, located on the α1 helix of the DP α-chain, leads to a difference in the binding affinity with peptides derived from spike protein. The alanine of HLA-DPA1*01:03 α-chain had intermolecular contact with tyrosine in the peptide core, whereas the methionine of HLA-DPA1*03:01 α-chain prevented high-affinity binding with tyrosine due to steric clash. The latter may compromise the ability of HLA-DPA1*03:01 to present antigens optimally and activate the CD4^+^ T cells, thereby undermines the effector function, specifically its ability to clear the virus causing infection effectively. The data suggest that HLA-DPA1*01:03 may be protective because of effects on immunodomiant peptide binding, whereas HLA-DPA1*03:01 was associated with risk to SARS-CoV-2 infection because of limitations on binding immunodominant SARS-CoV-2 epitopes.

The limitations of this study were the small population cohort and the lack of patient clinical information that could be extrapolated to examine the associations of other clinical symptoms and HLAs. Overall, this study demonstrates HLA typing and in-silico structural modeling to identify susceptible and protective HLA alleles. This approach can potentially provide a genetic biomarker to determine if an individual is protected from the severity of the infection or if an individual is susceptible to the disease. These biomarkers may be essential in the decision-making process for developing and implementing a strategy to keep the individual safe if there is no vaccine or treatment available.

## Data Availability

The data that support the findings of this study are presented in the manuscript and available in the Supplementary Materials. The data can also be available from the corresponding author upon request

## Author Contributions

YS and SR conducted the experiments. PJ obtained patient samples and involved in the experimental design. YS, DO, and CN conceptualized the study, performed data analysis, and prepared the manuscript. All authors have read and approved the final manuscript.

## Acknowledgments

CQN is supported financially in part by PHS grants DE023433, DE018958, and DE028544 from the National Institutes of Health.

