## Supplemental Tables 1 & 2 & 3 for "Identification of risk and protective human leukocyte antigens in COVID-19 using genotyping and structural modeling"

**Table S1 Full list of alleles from genotyping: the number of alleles at the HLA-A, -B, -C, -DPA1, -DPB1, -DQA1, -DQB1, -DRB1 loci were 30, 46, 20, 5, 20, 7, 16, and 32, respectively.**

| Allele | COVID-19 | | Control group | | Odds ratio (95% CI) | P-value |
|---|---|---|---|---|---|---|
| | N | Frequency (%) | N | Frequency (%) | | |
| **A*01:01** | 17 | 10.3659 | 18 | 12.1622 | 0.8357(0.3933 - 1.7448) | 0.7200 |
| **A*02:01** | 29 | 17.6829 | 39 | 26.3514 | 0.6014(0.3418 - 1.0602) | 0.0745 |
| **A*02:02** | 2 | 1.2195 | 1 | 0.6757 | 1.8115(0.1405 - 52.9657) | 0.9288 |
| **A*02:05** | 1 | 0.6098 | 1 | 0.6757 | 0.9021(0.0233 - 34.9456) | 0.5238 |
| **A*02:06** | 0 | 0.0000 | 1 | 0.6757 | 0(0 - 17.1463) | 0.4744 |
| **A*03:01** | 17 | 10.3659 | 16 | 10.8108 | 0.9542(0.437 - 2.0133) | 0.9548 |
| **A*11:01** | 9 | 5.4878 | 11 | 7.4324 | 0.7239(0.2631 - 1.8442) | 0.4983 |
| **A*23:01** | 9 | 5.4878 | 4 | 2.7027 | 2.0856(0.6398 - 7.4491) | 0.2651 |
| **A*24:02** | 15 | 9.1463 | 10 | 6.7568 | 1.3878(0.5683 - 3.2532) | 0.5327 |
| **A*25:01** | 3 | 1.8293 | 3 | 2.0270 | 0.9009(0.1606 - 5.0545) | 0.7751 |
| **A*26:01** | 4 | 2.4390 | 6 | 4.0541 | 0.5927(0.154 - 2.3371) | 0.5262 |
| **A*26:08** | 0 | 0.0000 | 1 | 0.6757 | 0(0 - 17.1463) | 0.4744 |
| **A*29:01** | 0 | 0.0000 | 1 | 0.6757 | 0(0 - 17.1463) | 0.4744 |
| **A*29:02** | 4 | 2.4390 | 7 | 4.7297 | 0.5047(0.1353 - 1.7824) | 0.3609 |
| **A*30:01** | 8 | 4.8780 | 1 | 0.6757 | 7.5017(1.1526 - 165.9402) | 0.0386 |
| **A*30:02** | 7 | 4.2683 | 3 | 2.0270 | 2.15(0.5601 - 9.805) | 0.3424 |
| **A*31:01** | 2 | 1.2195 | 4 | 2.7027 | 0.4456(0.059 - 2.4295) | 0.4281 |
| **A*32:01** | 4 | 2.4390 | 0 | 0.0000 | Inf(0.8162 - Inf) | 0.1246 |
| **A*33:01** | 2 | 1.2195 | 4 | 2.7027 | 0.4456(0.059 - 2.4295) | 0.4281 |
| **A*33:03** | 3 | 1.8293 | 3 | 2.0270 | 0.9009(0.1606 - 5.0545) | 0.7751 |
| **A*34:01** | 2 | 1.2195 | 0 | 0.0000 | Inf(0.2602 - Inf) | 0.4997 |
| **A*34:02** | 4 | 2.4390 | 1 | 0.6757 | 3.6616(0.4708 - 89.6824) | 0.3743 |
| **A*36:01** | 3 | 1.8293 | 1 | 0.6757 | 2.7309(0.2978 - 71.21) | 0.6246 |
| **A*66:01** | 2 | 1.2195 | 0 | 0.0000 | Inf(0.2602 - Inf) | 0.4997 |
| **A*66:02** | 1 | 0.6098 | 0 | 0.0000 | Inf(0.0475 - Inf) | 0.9590 |
| **A*68:01** | 4 | 2.4390 | 8 | 5.4054 | 0.4386(0.1205 - 1.5882) | 0.2400 |
| **A*68:02** | 2 | 1.2195 | 2 | 1.3514 | 0.9015(0.0964 - 8.4273) | 0.6888 |
| **A*68:17** | 0 | 0.0000 | 1 | 0.6757 | 0(0 - 17.1463) | 0.4744 |

| | | | | | | |
|---|---|---|---|---|---|---|
| **A*74:01** | 9 | 5.4878 | 1 | 0.6757 | 8.4924(1.2738 - 185.6196) | 0.0211 |
| **A*80:01** | 1 | 0.6098 | 0 | 0.0000 | Inf(0.0475 - Inf) | 0.9590 |
| **B*07:02** | 13 | 9.5588 | 9 | 7.7586 | 1.2554(0.4989 - 3.363) | 0.6603 |
| **B*07:05** | 1 | 0.7353 | 2 | 1.7241 | 0.4236(0.0145 - 5.4739) | 0.5959 |
| **B*08:01** | 13 | 9.5588 | 12 | 10.3448 | 0.9163(0.3955 - 2.195) | 0.8363 |
| **B*13:02** | 2 | 1.4706 | 2 | 1.7241 | 0.8513(0.0908 - 7.9793) | 0.7300 |
| **B*14:01** | 1 | 0.7353 | 4 | 3.4483 | 0.2085(0.0085 - 1.6275) | 0.1836 |
| **B*14:02** | 6 | 4.4118 | 4 | 3.4483 | 1.291(0.3243 - 4.992) | 0.7569 |
| **B*15:01** | 8 | 5.8824 | 5 | 4.3103 | 1.3857(0.4156 - 4.4814) | 0.7765 |
| **B*15:03** | 6 | 4.4118 | 1 | 0.8621 | 5.2797(0.7152 - 121.2847) | 0.1281 |
| **B*15:10** | 0 | 0.0000 | 1 | 0.8621 | 0(0 - 16.2059) | 0.4603 |
| **B*15:16** | 1 | 0.7353 | 2 | 1.7241 | 0.4236(0.0145 - 5.4739) | 0.5959 |
| **B*15:17** | 0 | 0.0000 | 2 | 1.7241 | 0(0 - 2.9543) | 0.2109 |
| **B*15:26** | 1 | 0.7353 | 0 | 0.0000 | Inf(0.0449 - Inf) | 0.9364 |
| **B*18:01** | 5 | 3.6765 | 3 | 2.5862 | 1.4356(0.3428 - 7.0318) | 0.7292 |
| **B*27:05** | 3 | 2.2059 | 8 | 6.8966 | 0.3059(0.0683 - 1.2853) | 0.1186 |
| **B*35:01** | 6 | 4.4118 | 5 | 4.3103 | 1.0245(0.2807 - 3.513) | 0.7871 |
| **B*35:02** | 2 | 1.4706 | 1 | 0.8621 | 1.7129(0.1326 - 50.1855) | 0.8897 |
| **B*35:03** | 1 | 0.7353 | 2 | 1.7241 | 0.4236(0.0145 - 5.4739) | 0.5959 |
| **B*37:01** | 1 | 0.7353 | 1 | 0.8621 | 0.8524(0.022 - 33.0693) | 0.5491 |
| **B*38:01** | 1 | 0.7353 | 1 | 0.8621 | 0.8524(0.022 - 33.0693) | 0.5491 |
| **B*38:02** | 1 | 0.7353 | 0 | 0.0000 | Inf(0.0449 - Inf) | 0.9364 |
| **B*39:01** | 4 | 2.9412 | 2 | 1.7241 | 1.7237(0.3149 - 13.0633) | 0.6898 |
| **B*39:06** | 1 | 0.7353 | 0 | 0.0000 | Inf(0.0449 - Inf) | 0.9364 |
| **B*39:10** | 1 | 0.7353 | 0 | 0.0000 | Inf(0.0449 - Inf) | 0.9364 |
| **B*40:01** | 5 | 3.6765 | 8 | 6.8966 | 0.5166(0.1597 - 1.7222) | 0.2688 |
| **B*40:02** | 2 | 1.4706 | 1 | 0.8621 | 1.7129(0.1326 - 50.1855) | 0.8897 |
| **B*40:06** | 0 | 0.0000 | 1 | 0.8621 | 0(0 - 16.2059) | 0.4603 |
| **B*41:01** | 0 | 0.0000 | 3 | 2.5862 | 0(0 - 1.4517) | 0.0962 |
| **B*42:01** | 6 | 4.4118 | 0 | 0.0000 | 11.6054(0.6467 - 208.2604) | 0.0323 |

| | | | | | | |
|---|---|---|---|---|---|---|
| **B*44:02** | 5 | 3.6765 | 10 | 8.6207 | 0.406(0.1303 - 1.3326) | 0.1145 |
| **B*44:03** | 8 | 5.8824 | 5 | 4.3103 | 1.3857(0.4156 - 4.4814) | 0.7765 |
| **B*45:01** | 2 | 1.4706 | 3 | 2.5862 | 0.5635(0.0689 - 3.6814) | 0.6639 |
| **B*47:01** | 1 | 0.7353 | 0 | 0.0000 | Inf(0.0449 - Inf) | 0.9364 |
| **B*49:01** | 5 | 3.6765 | 0 | 0.0000 | Inf(0.8507 - Inf) | 0.0639 |
| **B*50:01** | 3 | 2.2059 | 0 | 0.0000 | Inf(0.4998 - Inf) | 0.2518 |
| **B*51:01** | 0 | 0.0000 | 5 | 4.3103 | 0.0743(0.0041 - 1.3576) | 0.0197 |
| **B*51:08** | 1 | 0.7353 | 0 | 0.0000 | Inf(0.0449 - Inf) | 0.9364 |
| **B*52:01** | 1 | 0.7353 | 2 | 1.7241 | 0.4236(0.0145 - 5.4739) | 0.5959 |
| **B*53:01** | 3 | 2.2059 | 2 | 1.7241 | 1.2844(0.1966 - 10.5059) | 0.8573 |
| **B*55:01** | 1 | 0.7353 | 2 | 1.7241 | 0.4236(0.0145 - 5.4739) | 0.5959 |
| **B*56:01** | 1 | 0.7353 | 1 | 0.8621 | 0.8524(0.022 - 33.0693) | 0.5491 |
| **B*57:01** | 4 | 2.9412 | 3 | 2.5862 | 1.1408(0.2438 - 5.9027) | 0.8308 |
| **B*57:02** | 1 | 0.7353 | 0 | 0.0000 | Inf(0.0449 - Inf) | 0.9364 |
| **B*57:03** | 3 | 2.2059 | 1 | 0.8621 | 2.585(0.2811 - 67.5593) | 0.6268 |
| **B*58:01** | 3 | 2.2059 | 0 | 0.0000 | Inf(0.4998 - Inf) | 0.2518 |
| **B*58:02** | 3 | 2.2059 | 1 | 0.8621 | 2.585(0.2811 - 67.5593) | 0.6268 |
| **B*81:01** | 0 | 0.0000 | 1 | 0.8621 | 0(0 - 16.2059) | 0.4603 |
| **C*01:02** | 7 | 4.6053 | 4 | 2.8571 | 1.6387(0.4637 - 6.1211) | 0.5449 |
| **C*02:02** | 7 | 4.6053 | 9 | 6.4286 | 0.7035(0.2169 - 2.053) | 0.6092 |
| **C*03:02** | 1 | 0.6579 | 0 | 0.0000 | Inf(0.0485 - Inf) | 0.9671 |
| **C*03:03** | 6 | 3.9474 | 4 | 2.8571 | 1.3957(0.353 - 5.3785) | 0.7518 |
| **C*03:04** | 13 | 8.5526 | 13 | 9.2857 | 0.914(0.4046 - 2.0649) | 0.8402 |
| **C*04:01** | 25 | 16.4474 | 16 | 11.4286 | 1.5234(0.7649 - 3.1659) | 0.2410 |
| **C*05:01** | 9 | 5.9211 | 14 | 10.0000 | 0.5675(0.2175 - 1.3523) | 0.2768 |
| **C*06:02** | 17 | 11.1842 | 11 | 7.8571 | 1.4748(0.6602 - 3.5313) | 0.4270 |
| **C*07:01** | 18 | 11.8421 | 17 | 12.1429 | 0.972(0.4626 - 2.0768) | 0.9193 |
| **C*07:02** | 19 | 12.5000 | 12 | 8.5714 | 1.5216(0.6783 - 3.3219) | 0.3427 |
| **C*07:04** | 0 | 0.0000 | 3 | 2.1429 | 0(0 - 1.5701) | 0.1090 |
| **C*08:02** | 9 | 5.9211 | 8 | 5.7143 | 1.0383(0.3533 - 2.8246) | 0.8613 |
| **C*12:02** | 0 | 0.0000 | 2 | 1.4286 | 0(0 - 3.1922) | 0.2290 |

| | | | | | | |
|---|---|---|---|---|---|---|
| **C*12:03** | 4 | 2.6316 | 3 | 2.1429 | 1.2333(0.2644 - 6.3638) | 0.9123 |
| **C*14:02** | 1 | 0.6579 | 4 | 2.8571 | 0.2262(0.0092 - 1.7608) | 0.1977 |
| **C*15:02** | 4 | 2.6316 | 4 | 2.8571 | 0.9192(0.2156 - 3.918) | 0.8097 |
| **C*16:01** | 4 | 2.6316 | 8 | 5.7143 | 0.4471(0.1226 - 1.624) | 0.2418 |
| **C*16:02** | 1 | 0.6579 | 2 | 1.4286 | 0.4581(0.0157 - 5.9103) | 0.6088 |
| **C*17:01** | 4 | 2.6316 | 3 | 2.1429 | 1.2333(0.2644 - 6.3638) | 0.9123 |
| **C*18:01** | 3 | 1.9737 | 3 | 2.1429 | 0.9197(0.1638 - 5.1651) | 0.7558 |
| **DPA1*01:03** | 101 | 60.1190 | 107 | 72.2973 | 0.5786(0.3551 - 0.9401) | 0.0244 |
| **DPA1*01:04** | 1 | 0.5952 | 1 | 0.6757 | 0.8806(0.0227 - 34.1086) | 0.5347 |
| **DPA1*02:01** | 39 | 23.2143 | 31 | 20.9459 | 1.1406(0.6558 - 1.9565) | 0.6846 |
| **DPA1*02:02** | 17 | 10.1190 | 8 | 5.4054 | 1.9661(0.7972 - 4.8757) | 0.1457 |
| **DPA1*03:01** | 10 | 5.9524 | 1 | 0.6757 | 9.2568(1.3264 - 200.3514) | 0.0120 |
| **DPB1*01:01** | 23 | 15.5405 | 8 | 5.7971 | 2.9792(1.2446 - 7.258) | 0.0123 |
| **DPB1*02:01** | 23 | 15.5405 | 19 | 13.7681 | 1.1518(0.5835 - 2.2667) | 0.7393 |
| **DPB1*03:01** | 8 | 5.4054 | 13 | 9.4203 | 0.5506(0.2161 - 1.4255) | 0.2571 |
| **DPB1*04:01** | 43 | 29.0541 | 53 | 38.4058 | 0.6578(0.3906 - 1.087) | 0.1042 |
| **DPB1*04:02** | 19 | 12.8378 | 10 | 7.2464 | 1.8812(0.8315 - 4.3458) | 0.1692 |
| **DPB1*05:01** | 3 | 2.0270 | 3 | 2.1739 | 0.9313(0.1658 - 5.2316) | 0.7442 |
| **DPB1*06:01** | 3 | 2.0270 | 3 | 2.1739 | 0.9313(0.1658 - 5.2316) | 0.7442 |
| **DPB1*09:01** | 2 | 1.3514 | 2 | 1.4493 | 0.9318(0.0996 - 8.7183) | 0.6647 |
| **DPB1*10:01** | 2 | 1.3514 | 5 | 3.6232 | 0.3656(0.0508 - 1.7964) | 0.2682 |
| **DPB1*11:01** | 2 | 1.3514 | 9 | 6.5217 | 0.1974(0.0301 - 0.9301) | 0.0301 |
| **DPB1*13:01** | 5 | 3.3784 | 1 | 0.7246 | 4.7683(0.6329 - 112.3687) | 0.2155 |
| **DPB1*14:01** | 2 | 1.3514 | 2 | 1.4493 | 0.9318(0.0996 - 8.7183) | 0.6647 |
| **DPB1*15:01** | 1 | 0.6757 | 1 | 0.7246 | 0.9322(0.0241 - 36.1305) | 0.5090 |
| **DPB1*16:01** | 1 | 0.6757 | 1 | 0.7246 | 0.9322(0.0241 - 36.1305) | 0.5090 |

| | | | | | | |
|---|---|---|---|---|---|---|
| **DPB1*17:01** | 5 | 3.3784 | 4 | 2.8986 | 1.1707(0.308 - 4.7058) | 0.9151 |
| **DPB1*18:01** | 1 | 0.6757 | 2 | 1.4493 | 0.4638(0.0159 - 5.9843) | 0.6109 |
| **DPB1*19:01** | 1 | 0.6757 | 2 | 1.4493 | 0.4638(0.0159 - 5.9843) | 0.6109 |
| **DPB1*26:01** | 1 | 0.6757 | 0 | 0.0000 | Inf(0.0491 - Inf) | 0.9720 |
| **DPB1*40:01** | 2 | 1.3514 | 0 | 0.0000 | Inf(0.2689 - Inf) | 0.4989 |
| **DPB1*85:01** | 1 | 0.6757 | 0 | 0.0000 | Inf(0.0491 - Inf) | 0.9720 |
| **DQA1*01:01** | 20 | 13.1579 | 22 | 15.9420 | 0.7995(0.3938 - 1.5854) | 0.5096 |
| **DQA1*01:02** | 42 | 27.6316 | 33 | 23.9130 | 1.2141(0.7066 - 2.121) | 0.5038 |
| **DQA1*01:03** | 8 | 5.2632 | 12 | 8.6957 | 0.5844(0.2273 - 1.5958) | 0.3537 |
| **DQA1*02:01** | 14 | 9.2105 | 26 | 18.8406 | 0.4383(0.2122 - 0.8942) | 0.0257 |
| **DQA1*03:01** | 22 | 14.4737 | 21 | 15.2174 | 0.9431(0.4802 - 1.8702) | 0.8702 |
| **DQA1*04:01** | 6 | 3.9474 | 2 | 1.4493 | 2.7858(0.4998 - 19.382) | 0.2869 |
| **DQA1*05:01** | 40 | 26.3158 | 22 | 15.9420 | 1.8791(1.0435 - 3.5288) | 0.0326 |
| **DQB1*02:01** | 18 | 11.2500 | 14 | 10.1449 | 1.1223(0.5099 - 2.4659) | 0.8519 |
| **DQB1*02:02** | 8 | 5.0000 | 14 | 10.1449 | 0.4674(0.185 - 1.1564) | 0.1194 |
| **DQB1*03:01** | 21 | 13.1250 | 18 | 13.0435 | 1.0072(0.4873 - 2.0192) | 0.8796 |
| **DQB1*03:02** | 13 | 8.1250 | 15 | 10.8696 | 0.726(0.3232 - 1.6491) | 0.4334 |
| **DQB1*03:03** | 8 | 5.0000 | 7 | 5.0725 | 0.985(0.3363 - 3.1954) | 0.8125 |
| **DQB1*03:19** | 7 | 4.3750 | 0 | 0.0000 | 13.5342(0.7659 - 239.1722) | 0.0163 |
| **DQB1*04:02** | 7 | 4.3750 | 3 | 2.1739 | 2.0541(0.5348 - 9.376) | 0.3491 |
| **DQB1*05:01** | 21 | 13.1250 | 23 | 16.6667 | 0.7561(0.3852 - 1.4543) | 0.4163 |
| **DQB1*05:02** | 4 | 2.5000 | 3 | 2.1739 | 1.1533(0.2473 - 5.9491) | 0.8429 |
| **DQB1*05:03** | 4 | 2.5000 | 4 | 2.8986 | 0.8594(0.2017 - 3.6621) | 0.8830 |
| **DQB1*05:04** | 1 | 0.6250 | 0 | 0.0000 | Inf(0.0454 - Inf) | 0.9409 |
| **DQB1*06:01** | 2 | 1.2500 | 2 | 1.4493 | 0.8612(0.0921 - 8.0547) | 0.7221 |
| **DQB1*06:02** | 30 | 18.7500 | 18 | 13.0435 | 1.5363(0.7976 - 2.9717) | 0.2077 |
| **DQB1*06:03** | 10 | 6.2500 | 9 | 6.5217 | 0.9557(0.3631 - 2.7037) | 0.8871 |
| **DQB1*06:04** | 3 | 1.8750 | 6 | 4.3478 | 0.4216(0.0897 - 1.8947) | 0.3112 |
| **DQB1*06:09** | 3 | 1.8750 | 2 | 1.4493 | 1.2982(0.1992 - 10.5931) | 0.8674 |

| | | | | | | |
|---|---|---|---|---|---|---|
| **DRB1*01:01** | 8 | 5.4795 | 13 | 10.0000 | 0.523(0.2051 - 1.3577) | 0.1777 |
| **DRB1*01:02** | 4 | 2.7397 | 1 | 0.7692 | 3.619(0.4645 - 88.7575) | 0.3747 |
| **DRB1*03:01** | 17 | 11.6438 | 10 | 7.6923 | 1.5788(0.6831 - 3.6726) | 0.3137 |
| **DRB1*03:02** | 1 | 0.6849 | 2 | 1.5385 | 0.4426(0.0151 - 5.7137) | 0.6030 |
| **DRB1*04:01** | 8 | 5.4795 | 9 | 6.9231 | 0.7801(0.2858 - 2.2825) | 0.6271 |
| **DRB1*04:02** | 1 | 0.6849 | 1 | 0.7692 | 0.89(0.023 - 34.5052) | 0.5297 |
| **DRB1*04:03** | 0 | 0.0000 | 1 | 0.7692 | 0(0 - 16.9178) | 0.4710 |
| **DRB1*04:04** | 3 | 2.0548 | 5 | 3.8462 | 0.5257(0.1075 - 2.1972) | 0.4815 |
| **DRB1*04:05** | 2 | 1.3699 | 0 | 0.0000 | Inf(0.2568 - Inf) | 0.4999 |
| **DRB1*04:07** | 0 | 0.0000 | 1 | 0.7692 | 0(0 - 16.9178) | 0.4710 |
| **DRB1*07:01** | 14 | 9.5890 | 23 | 17.6923 | 0.4947(0.2338 - 1.0588) | 0.0532 |
| **DRB1*08:01** | 2 | 1.3699 | 1 | 0.7692 | 1.788(0.1385 - 52.3379) | 0.9194 |
| **DRB1*08:04** | 1 | 0.6849 | 0 | 0.0000 | Inf(0.0469 - Inf) | 0.9536 |
| **DRB1*08:06** | 1 | 0.6849 | 0 | 0.0000 | Inf(0.0469 - Inf) | 0.9536 |
| **DRB1*09:01** | 2 | 1.3699 | 1 | 0.7692 | 1.788(0.1385 - 52.3379) | 0.9194 |
| **DRB1*10:01** | 3 | 2.0548 | 2 | 1.5385 | 1.3412(0.2056 - 10.9552) | 0.8957 |
| **DRB1*11:01** | 10 | 6.8493 | 9 | 6.9231 | 0.9886(0.3738 - 2.8097) | 0.8306 |
| **DRB1*11:02** | 4 | 2.7397 | 2 | 1.5385 | 1.7991(0.3293 - 13.6148) | 0.6873 |
| **DRB1*11:04** | 4 | 2.7397 | 2 | 1.5385 | 1.7991(0.3293 - 13.6148) | 0.6873 |
| **DRB1*12:01** | 1 | 0.6849 | 4 | 3.0769 | 0.2183(0.0089 - 1.701) | 0.1912 |
| **DRB1*13:01** | 15 | 10.2740 | 10 | 7.6923 | 1.3725(0.5565 - 3.2232) | 0.5314 |
| **DRB1*13:02** | 6 | 4.1096 | 10 | 7.6923 | Inf(0.5217 - Inf) | 0.2498 |
| **DRB1*13:03** | 3 | 2.0548 | 0 | 0.0000 | 0.5155(0.1738 - 1.563) | 0.3021 |
| **DRB1*13:04** | 5 | 3.4247 | 0 | 0.0000 | Inf(0.8889 - Inf) | 0.0623 |
| **DRB1*14:01** | 2 | 1.3699 | 2 | 1.5385 | 0.8893(0.095 - 8.3249) | 0.6984 |
| **DRB1*14:02** | 2 | 1.3699 | 0 | 0.0000 | Inf(0.2568 - Inf) | 0.4999 |
| **DRB1*14:03** | 1 | 0.6849 | 0 | 0.0000 | Inf(0.0469 - Inf) | 0.9536 |
| **DRB1*15:01** | 13 | 8.9041 | 12 | 9.2308 | 0.9613(0.4181 - 2.2854) | 0.9079 |
| **DRB1*15:02** | 2 | 1.3699 | 2 | 1.5385 | 0.8893(0.095 - 8.3249) | 0.6984 |
| **DRB1*15:03** | 9 | 6.1644 | 4 | 3.0769 | 2.0642(0.6294 - 7.3926) | 0.2661 |
| **DRB1*16:01** | 1 | 0.6849 | 3 | 2.3077 | 0.2932(0.0112 - 2.6928) | 0.3456 |

| | | | | | | |
|---|---|---|---|---|---|---|
| **DRB1*16:02** | 1 | 0.6849 | 0 | 0.0000 | Inf(0.0469 - Inf) | 0.9536 |

**Table S2 High rates of ambiguous imputation in HLA-DRB/3/4/5.**

| Alleles | Frequency (%) |
|---|---|
| **DRB3*01:01** | 18.1818 |
| **DRB3*02:02** | 22.0779 |
| **DRB3*02:10** | 0.6494 |
| **DRB3*03:01** | 7.1429 |
| **DRB3*undefined** | 51.9481 |
| **DRB4*01:01** | 5.4217 |
| **DRB4*01:03** | 16.2651 |
| **DRB4*undefined** | 78.3133 |
| **DRB5*01:01** | 13.6905 |
| **DRB5*01:02** | 0.5952 |
| **DRB5*02:02** | 1.7857 |
| **DRB5*undefined** | 83.9286 |

**Table S3 Summary of alleles genotyping in the black population: the number of alleles at the HLA-A, -B, -C, -DPA1, -DPB1, -DQA1, -DQB1, -DRB1 loci were 19, 18,17, 4, 12, 6, 11 and 19, respectively.**

| Allele | COVID-19 | | Control group | | Odds ratio (95% CI) | P-value |
|---|---|---|---|---|---|---|
| | N | Frequency (%) | N | Frequency (%) | | |
| **A*01:01** | 3 | 7.8947 | 1 | 7.1429 | 1.112(0.1117-30.936) | 1 |
| **A*02:01** | 1 | 2.6316 | 4 | 28.5714 | 0.0722(0.0027-0.6233) | 0.0154 |
| **A*02:02** | 1 | 2.6316 | 0 | 0.0000 | Inf(0.0194-Inf) | 1 |
| **A*03:01** | 2 | 5.2632 | 1 | 7.1429 | 0.7271(0.0529-22.5142) | 1 |
| **A*11:01** | 1 | 2.6316 | 0 | 0.0000 | Inf(0.0194-Inf) | 1 |
| **A*23:01** | 3 | 7.8947 | 1 | 7.1429 | 1.112(0.1117-30.936) | 1 |
| **A*24:02** | 1 | 2.6316 | 0 | 0.0000 | Inf(0.0194-Inf) | 1 |
| **A*26:01** | 1 | 2.6316 | 0 | 0.0000 | Inf(0.0194-Inf) | 1 |
| **A*29:02** | 1 | 2.6316 | 2 | 14.2857 | 0.1693(0.0055-2.3288) | 0.1729 |
| **A*30:01** | 5 | 13.1579 | 0 | 0.0000 | Inf(0.3866-Inf) | 0.307 |
| **A*30:02** | 3 | 7.8947 | 0 | 0.0000 | Inf(0.2129-Inf) | 0.5547 |
| **A*33:01** | 0 | 0.0000 | 2 | 14.2857 | 0(0-1.2349) | 0.0686 |
| **A*33:03** | 3 | 7.8947 | 1 | 7.1429 | 1.112(0.1117-30.936) | 1 |
| **A*34:02** | 2 | 5.2632 | 1 | 7.1429 | 0.7271(0.0529-22.5142) | 1 |
| **A*36:01** | 2 | 5.2632 | 0 | 0.0000 | Inf(0.1048-Inf) | 1 |
| **A*66:01** | 2 | 5.2632 | 0 | 0.0000 | Inf(0.1048-Inf) | 1 |
| **A*68:01** | 1 | 2.6316 | 1 | 7.1429 | 0.3598(0.0089-14.5474) | 0.4698 |
| **A*68:02** | 1 | 2.6316 | 0 | 0.0000 | Inf(0.0194-Inf) | 1 |
| **A*74:01** | 5 | 13.1579 | 0 | 0.0000 | Inf(0.3866-Inf) | 0.307 |
| **B*07:02** | 1 | 3.5714 | 0 | 0.0000 | Inf(0.0113-Inf) | 1 |
| **B*08:01** | 4 | 14.2857 | 1 | 16.6667 | 0.838(0.0823-24.146) | 1 |
| **B*14:02** | 2 | 7.1429 | 0 | 0.0000 | Inf(0.0589-Inf) | 1 |
| **B*15:03** | 4 | 14.2857 | 0 | 0.0000 | Inf(0.1645-Inf) | 0.5762 |
| **B*15:16** | 1 | 3.5714 | 1 | 16.6667 | 0.1992(0.0047-8.5328) | 0.3262 |
| **B*15:17** | 0 | 0.0000 | 1 | 16.6667 | 0(0-4.0714) | 0.1765 |
| **B*27:05** | 0 | 0.0000 | 1 | 16.6667 | 0(0-4.0714) | 0.1765 |
| **B*35:01** | 1 | 3.5714 | 1 | 16.6667 | 0.1992(0.0047-8.5328) | 0.3262 |
| **B*39:10** | 1 | 3.5714 | 0 | 0.0000 | Inf(0.0113-Inf) | 1 |
| **B*42:01** | 2 | 7.1429 | 0 | 0.0000 | Inf(0.0589-Inf) | 1 |
| **B*44:03** | 1 | 3.5714 | 0 | 0.0000 | Inf(0.0113-Inf) | 1 |
| **B*45:01** | 1 | 3.5714 | 0 | 0.0000 | Inf(0.0113-Inf) | 1 |
| **B*49:01** | 1 | 3.5714 | 0 | 0.0000 | Inf(0.0113-Inf) | 1 |
| **B*50:01** | 1 | 3.5714 | 0 | 0.0000 | Inf(0.0113-Inf) | 1 |
| **B*53:01** | 0 | 0.0000 | 1 | 16.6667 | 0(0-4.0714) | 0.1765 |
| **B*57:03** | 2 | 7.1429 | 0 | 0.0000 | Inf(0.0589-Inf) | 1 |
| **B*58:01** | 3 | 10.7143 | 0 | 0.0000 | Inf(0.1173-Inf) | 1 |
| **B*58:02** | 3 | 10.7143 | 0 | 0.0000 | Inf(0.1173-Inf) | 1 |
| **C*01:02** | 2 | 6.6667 | 1 | 7.1429 | 0.9302(0.0673-28.9434) | 1 |
| **C*02:02** | 0 | 0.0000 | 1 | 7.1429 | 0(0-8.8667) | 0.3182 |
| **C*03:02** | 1 | 3.3333 | 0 | 0.0000 | Inf(0.0246-Inf) | 1 |

| | | | | | | |
|---|---|---|---|---|---|---|
| **C*03:04** | 2 | 6.6667 | 0 | 0.0000 | Inf(0.1334-Inf) | 0.556 |
| **C*04:01** | 4 | 13.3333 | 4 | 28.5714 | 0.394(0.0786-1.9557) | 0.4024 |
| **C*05:01** | 0 | 0.0000 | 1 | 7.1429 | 0(0-8.8667) | 0.3182 |
| **C*06:02** | 7 | 23.3333 | 1 | 7.1429 | 3.8567(0.4904-94.3989) | 0.2477 |
| **C*07:01** | 4 | 13.3333 | 2 | 14.2857 | 0.9248(0.1496-7.7518) | 1 |
| **C*07:02** | 2 | 6.6667 | 0 | 0.0000 | Inf(0.1334-Inf) | 0.556 |
| **C*08:02** | 3 | 10.0000 | 0 | 0.0000 | Inf(0.2724-Inf) | 0.5402 |
| **C*12:03** | 1 | 3.3333 | 0 | 0.0000 | Inf(0.0246-Inf) | 1 |
| **C*14:02** | 1 | 3.3333 | 1 | 7.1429 | 0.4574(0.0113-18.559) | 1 |
| **C*15:02** | 0 | 0.0000 | 1 | 7.1429 | 0(0-8.8667) | 0.3182 |
| **C*16:01** | 1 | 3.3333 | 0 | 0.0000 | Inf(0.0246-Inf) | 1 |
| **C*16:02** | 0 | 0.0000 | 1 | 7.1429 | 0(0-8.8667) | 0.3182 |
| **C*17:01** | 1 | 3.3333 | 1 | 7.1429 | 0.4574(0.0113-18.559) | 1 |
| **C*18:01** | 1 | 3.3333 | 0 | 0.0000 | Inf(0.0246-Inf) | 1 |
| **DPA1*01:03** | 17 | 44.7368 | 6 | 42.8571 | 1.0778(0.2999-4.3623) | 1 |
| **DPA1*02:01** | 13 | 34.2105 | 9 | 64.2857 | 0.2963(0.08-1.2079) | 0.0647 |
| **DPA1*02:02** | 3 | 7.8947 | 0 | 0.0000 | Inf(0.2129-Inf) | 0.5547 |
| **DPA1*03:01** | 5 | 13.1579 | 1 | 7.1429 | 1.9474(0.2086-49.3099) | 0.6701 |
| **DPB1*01:01** | 6 | 17.6471 | 4 | 28.5714 | 0.5433(0.1203-2.4539) | 0.448 |
| **DPB1*02:01** | 7 | 20.5882 | 2 | 14.2857 | 1.5422(0.2417-11.6885) | 0.7081 |
| **DPB1*03:01** | 0 | 0.0000 | 1 | 7.1429 | 0(0-7.8235) | 0.2917 |
| **DPB1*04:01** | 4 | 11.7647 | 2 | 14.2857 | 0.8039(0.1307-6.7057) | 1 |
| **DPB1*04:02** | 5 | 14.7059 | 0 | 0.0000 | Inf(0.44-Inf) | 0.3028 |
| **DPB1*11:01** | 1 | 2.9412 | 2 | 14.2857 | 0.1896(0.0061-2.6143) | 0.1999 |
| **DPB1*13:01** | 2 | 5.8824 | 0 | 0.0000 | Inf(0.1174-Inf) | 0.578 |
| **DPB1*14:01** | 1 | 2.9412 | 1 | 7.1429 | 0.4028(0.0099-16.3102) | 1 |
| **DPB1*17:01** | 4 | 11.7647 | 1 | 7.1429 | 1.7155(0.1996-45.1635) | 1 |
| **DPB1*18:01** | 1 | 2.9412 | 1 | 7.1429 | 0.4028(0.0099-16.3102) | 1 |
| **DPB1*40:01** | 1 | 2.9412 | 0 | 0.0000 | Inf(0.0217-Inf) | 1 |
| **DPB1*85:01** | 2 | 5.8824 | 0 | 0.0000 | Inf(0.1174-Inf) | 0.578 |
| **DQA1*01:01** | 4 | 12.5000 | 3 | 21.4286 | 0.5318(0.0991-3.1105) | 0.6576 |
| **DQA1*01:02** | 15 | 46.8750 | 4 | 28.5714 | 2.169(0.5148-8.9474) | 0.3352 |
| **DQA1*02:01** | 0 | 0.0000 | 1 | 7.1429 | 0(0-8.3125) | 0.3043 |
| **DQA1*03:01** | 2 | 6.2500 | 1 | 7.1429 | 0.8694(0.063-27.0148) | 1 |
| **DQA1*04:01** | 0 | 0.0000 | 1 | 7.1429 | 0(0-8.3125) | 0.3043 |
| **DQA1*05:01** | 11 | 34.3750 | 4 | 28.5714 | 1.302(0.3281-5.4555) | 0.7476 |
| **DQB1*02:01** | 5 | 13.8889 | 3 | 13.8889 | DQB1*02:01(5-13.8889) | 0.3939 |
| **DQB1*02:02** | 0 | 0.0000 | 1 | 0.0000 | DQB1*02:02(0-0) | 0.25 |
| **DQB1*03:01** | 4 | 11.1111 | 1 | 11.1111 | DQB1*03:01(4-11.1111) | 1 |
| **DQB1*03:02** | 1 | 2.7778 | 0 | 2.7778 | DQB1*03:02(1-2.7778) | 1 |
| **DQB1*03:03** | 1 | 2.7778 | 0 | 2.7778 | DQB1*03:03(1-2.7778) | 1 |
| **DQB1*03:19** | 3 | 8.3333 | 0 | 8.3333 | DQB1*03:19(3-8.3333) | 0.5629 |
| **DQB1*04:02** | 0 | 0.0000 | 1 | 0.0000 | DQB1*04:02(0-0) | 0.25 |

| | | | | | | |
|---|---|---|---|---|---|---|
| **DQB1*05:01** | 7 | 19.4444 | 4 | 19.4444 | DQB1*05:01(7-19.4444) | 0.4304 |
| **DQB1*06:02** | 13 | 36.1111 | 2 | 36.1111 | DQB1*06:02(13-36.1111) | 0.2919 |
| **DQB1*06:03** | 1 | 2.7778 | 0 | 2.7778 | DQB1*06:03(1-2.7778) | 1 |
| **DQB1*06:09** | 1 | 2.7778 | 0 | 2.7778 | DQB1*06:09(1-2.7778) | 1 |
| **DRB1*01:01** | 1 | 2.9412 | 1 | 8.3333 | 0.343(0.0084-13.9781) | 0.458 |
| **DRB1*01:02** | 1 | 2.9412 | 0 | 0.0000 | Inf(0.0186-Inf) | 1 |
| **DRB1*03:01** | 4 | 11.7647 | 3 | 25.0000 | 0.4093(0.0745-2.4367) | 0.3554 |
| **DRB1*03:02** | 0 | 0.0000 | 1 | 8.3333 | 0(0-6.7059) | 0.2609 |
| **DRB1*04:03** | 0 | 0.0000 | 1 | 8.3333 | 0(0-6.7059) | 0.2609 |
| **DRB1*08:06** | 1 | 2.9412 | 0 | 0.0000 | Inf(0.0186-Inf) | 1 |
| **DRB1*10:01** | 1 | 2.9412 | 0 | 0.0000 | Inf(0.0186-Inf) | 1 |
| **DRB1*11:01** | 3 | 8.8235 | 3 | 25.0000 | 0.2999(0.0463-1.9261) | 0.3167 |
| **DRB1*11:02** | 1 | 2.9412 | 0 | 0.0000 | Inf(0.0186-Inf) | 1 |
| **DRB1*12:01** | 0 | 0.0000 | 1 | 8.3333 | 0(0-6.7059) | 0.2609 |
| **DRB1*13:01** | 6 | 17.6471 | 0 | 0.0000 | Inf(0.464-Inf) | 0.1762 |
| **DRB1*13:02** | 1 | 2.9412 | 0 | 0.0000 | Inf(0.0186-Inf) | 1 |
| **DRB1*13:03** | 1 | 2.9412 | 0 | 0.0000 | Inf(0.0186-Inf) | 1 |
| **DRB1*13:04** | 5 | 14.7059 | 0 | 0.0000 | Inf(0.3836-Inf) | 0.3059 |
| **DRB1*14:01** | 1 | 2.9412 | 0 | 0.0000 | Inf(0.0186-Inf) | 1 |
| **DRB1*14:03** | 1 | 2.9412 | 0 | 0.0000 | Inf(0.0186-Inf) | 1 |
| **DRB1*15:01** | 1 | 2.9412 | 0 | 0.0000 | Inf(0.0186-Inf) | 1 |
| **DRB1*15:03** | 6 | 17.6471 | 1 | 8.3333 | 2.3201(0.2866-58.1266) | 0.6569 |
| **DRB1*16:01** | 0 | 0.0000 | 1 | 8.3333 | 0(0-6.7059) | 0.2609 |

**Table S4 Summary of alleles genotyping in the white population: the number of alleles at the HLA-A, -B, -C, -DPA1, -DPB1, -DQA1, -DQB1, -DRB1 loci were 21, 30, 18, 4, 15, 7, 15 and 20, respectively.**

| Allele | COVID-19 | | Control group | | Odds ratio (95% CI) | P-value |
|---|---|---|---|---|---|---|
| | N | Frequency (%) | N | Frequency (%) | | |
| **A*01:01** | 4 | 11.7647 | 10 | 11.6279 | 1.0132(0.2745-3.4462) | 1.0000 |
| **A*02:01** | 10 | 29.4118 | 23 | 26.7442 | 1.14(0.4528-2.8593) | 0.8219 |
| **A*02:05** | 0 | 0.0000 | 1 | 1.1628 | 0(0-48.0588) | 1.0000 |
| **A*02:06** | 0 | 0.0000 | 1 | 1.1628 | 0(0-48.0588) | 1.0000 |
| **A*03:01** | 6 | 17.6471 | 10 | 11.6279 | 1.6215(0.4988-4.9959) | 0.3845 |
| **A*11:01** | 1 | 2.9412 | 9 | 10.4651 | 0.2614(0.0117-1.7219) | 0.2787 |
| **A*23:01** | 1 | 2.9412 | 0 | 0.0000 | Inf(0.1331-Inf) | 0.2833 |
| **A*24:02** | 5 | 14.7059 | 7 | 8.1395 | 1.934(0.5567-6.6538) | 0.3172 |
| **A*25:01** | 0 | 0.0000 | 3 | 3.4884 | 0(0-4.3482) | 0.5575 |
| **A*26:01** | 1 | 2.9412 | 4 | 4.6512 | 0.6234(0.0249-4.9971) | 1.0000 |
| **A*26:08** | 0 | 0.0000 | 1 | 1.1628 | 0(0-48.0588) | 1.0000 |
| **A*29:01** | 0 | 0.0000 | 1 | 1.1628 | 0(0-48.0588) | 1.0000 |
| **A*29:02** | 1 | 2.9412 | 4 | 4.6512 | 0.6234(0.0249-4.9971) | 1.0000 |
| **A*30:01** | 0 | 0.0000 | 1 | 1.1628 | 0(0-48.0588) | 1.0000 |
| **A*30:02** | 0 | 0.0000 | 1 | 1.1628 | 0(0-48.0588) | 1.0000 |
| **A*31:01** | 2 | 5.8824 | 4 | 4.6512 | 1.2785(0.1643-7.2204) | 1.0000 |
| **A*32:01** | 1 | 2.9412 | 0 | 0.0000 | Inf(0.1331-Inf) | 0.2833 |
| **A*33:01** | 0 | 0.0000 | 1 | 1.1628 | 0(0-48.0588) | 1.0000 |
| **A*33:03** | 0 | 0.0000 | 1 | 1.1628 | 0(0-48.0588) | 1.0000 |
| **A*68:01** | 2 | 5.8824 | 3 | 3.4884 | 1.7205(0.2052-11.5549) | 0.6211 |
| **A*74:01** | 0 | 0.0000 | 1 | 1.1628 | 0(0-48.0588) | 1.0000 |
| **B*07:02** | 3 | 10.0000 | 6 | 8.5714 | 1.1831(0.2401-5.94) | 1.0000 |
| **B*07:05** | 0 | 0.0000 | 1 | 1.4286 | 0(0-44.3333) | 1.0000 |
| **B*08:01** | 2 | 6.6667 | 7 | 10.0000 | 0.6455(0.0914-3.3204) | 0.7202 |
| **B*13:02** | 1 | 3.3333 | 2 | 2.8571 | 1.1705(0.0391-15.4767) | 1.0000 |
| **B*14:01** | 0 | 0.0000 | 3 | 4.2857 | 0(0-4.0063) | 0.5519 |
| **B*14:02** | 1 | 3.3333 | 3 | 4.2857 | 0.7721(0.0288-7.3114) | 1.0000 |
| **B*15:01** | 2 | 6.6667 | 5 | 7.1429 | 0.9293(0.1243-4.9592) | 1.0000 |
| **B*15:10** | 0 | 0.0000 | 1 | 1.4286 | 0(0-44.3333) | 1.0000 |
| **B*18:01** | 1 | 3.3333 | 2 | 2.8571 | 1.1705(0.0391-15.4767) | 1.0000 |
| **B*27:05** | 1 | 3.3333 | 6 | 8.5714 | 0.3708(0.0157-2.7455) | 0.4386 |
| **B*35:01** | 1 | 3.3333 | 3 | 4.2857 | 0.7721(0.0288-7.3114) | 1.0000 |
| **B*35:02** | 1 | 3.3333 | 0 | 0.0000 | Inf(0.1228-Inf) | 0.3000 |
| **B*35:03** | 1 | 3.3333 | 2 | 2.8571 | 1.1705(0.0391-15.4767) | 1.0000 |
| **B*37:01** | 0 | 0.0000 | 1 | 1.4286 | 0(0-44.3333) | 1.0000 |
| **B*39:01** | 2 | 6.6667 | 1 | 1.4286 | 4.8392(0.3659-144.8529) | 0.2134 |
| **B*40:01** | 1 | 3.3333 | 4 | 5.7143 | 0.5718(0.0227-4.6168) | 1.0000 |
| **B*40:02** | 0 | 0.0000 | 1 | 1.4286 | 0(0-44.3333) | 1.0000 |
| **B*40:06** | 0 | 0.0000 | 1 | 1.4286 | 0(0-44.3333) | 1.0000 |

| | | | | | | |
|---|---|---|---|---|---|---|
| **B*41:01** | 0 | 0.0000 | 2 | 2.8571 | 0(0-8.1365) | 0.5758 |
| **B*44:02** | 4 | 13.3333 | 6 | 8.5714 | 1.6323(0.4011-7.2459) | 0.4816 |
| **B*44:03** | 3 | 10.0000 | 3 | 4.2857 | 2.4562(0.4197-14.3849) | 0.3608 |
| **B*45:01** | 0 | 0.0000 | 2 | 2.8571 | 0(0-8.1365) | 0.5758 |
| **B*49:01** | 1 | 3.3333 | 0 | 0.0000 | Inf(0.1228-Inf) | 0.3000 |
| **B*50:01** | 2 | 6.6667 | 0 | 0.0000 | Inf(0.6823-Inf) | 0.0879 |
| **B*51:01** | 0 | 0.0000 | 2 | 2.8571 | 0(0-8.1365) | 0.5758 |
| **B*52:01** | 0 | 0.0000 | 1 | 1.4286 | 0(0-44.3333) | 1.0000 |
| **B*55:01** | 0 | 0.0000 | 2 | 2.8571 | 0(0-8.1365) | 0.5758 |
| **B*56:01** | 0 | 0.0000 | 1 | 1.4286 | 0(0-44.3333) | 1.0000 |
| **B*57:01** | 2 | 6.6667 | 2 | 2.8571 | 2.4044(0.2493-23.1959) | 0.5812 |
| **B*57:02** | 1 | 3.3333 | 0 | 0.0000 | Inf(0.1228-Inf) | 0.3000 |
| **C*01:02** | 3 | 8.8235 | 3 | 3.7500 | 2.4616(0.4237-14.3103) | 0.3608 |
| **C*02:02** | 0 | 0.0000 | 6 | 7.5000 | 0(0-1.6512) | 0.1765 |
| **C*03:03** | 0 | 0.0000 | 4 | 5.0000 | 0(0-2.6108) | 0.3157 |
| **C*03:04** | 2 | 5.8824 | 9 | 11.2500 | 0.4957(0.0731-2.5922) | 0.5015 |
| **C*04:01** | 5 | 14.7059 | 6 | 7.5000 | 2.1109(0.5878-8.4442) | 0.2993 |
| **C*05:01** | 5 | 14.7059 | 8 | 10.0000 | 1.5454(0.4551-5.7532) | 0.5245 |
| **C*06:02** | 5 | 14.7059 | 9 | 11.2500 | 1.3564(0.4073-4.5498) | 0.7558 |
| **C*07:01** | 4 | 11.7647 | 9 | 11.2500 | 1.0514(0.2802-3.8546) | 1.0000 |
| **C*07:02** | 6 | 17.6471 | 9 | 11.2500 | 1.6821(0.5035-5.2972) | 0.3741 |
| **C*07:04** | 0 | 0.0000 | 1 | 1.2500 | 0(0-44.7059) | 1.0000 |
| **C*08:02** | 1 | 2.9412 | 6 | 7.5000 | 0.3764(0.016-2.7808) | 0.4422 |
| **C*12:02** | 0 | 0.0000 | 1 | 1.2500 | 0(0-44.7059) | 1.0000 |
| **C*12:03** | 0 | 0.0000 | 1 | 1.2500 | 0(0-44.7059) | 1.0000 |
| **C*15:02** | 0 | 0.0000 | 1 | 1.2500 | 0(0-44.7059) | 1.0000 |
| **C*16:01** | 2 | 5.8824 | 4 | 5.0000 | 1.1856(0.1522-6.7034) | 1.0000 |
| **C*16:02** | 0 | 0.0000 | 1 | 1.2500 | 0(0-44.7059) | 1.0000 |
| **C*17:01** | 0 | 0.0000 | 1 | 1.2500 | 0(0-44.7059) | 1.0000 |
| **C*18:01** | 1 | 2.9412 | 1 | 1.2500 | 2.3734(0.0604-93.2687) | 1.0000 |
| **DPA1*01:03** | 27 | 79.4118 | 66 | 76.7442 | 1.1673(0.4349-3.6294) | 0.8133 |
| **DPA1*01:04** | 0 | 0.0000 | 1 | 1.1628 | 0(0-48.0588) | 1.0000 |
| **DPA1*02:01** | 4 | 11.7647 | 13 | 15.1163 | 0.7504(0.2099-2.5997) | 0.7760 |
| **DPA1*02:02** | 3 | 8.8235 | 6 | 6.9767 | 1.2875(0.2638-6.3185) | 0.7117 |
| **DPB1*01:01** | 1 | 3.5714 | 2 | 2.5641 | 1.4025(0.0468-18.5584) | 1.0000 |
| **DPB1*02:01** | 3 | 10.7143 | 6 | 7.6923 | 1.4347(0.2908-7.2372) | 0.6959 |
| **DPB1*03:01** | 3 | 10.7143 | 6 | 7.6923 | 1.4347(0.2908-7.2372) | 0.6959 |
| **DPB1*04:01** | 11 | 39.2857 | 38 | 48.7179 | 0.6836(0.2792-1.6474) | 0.5081 |
| **DPB1*04:02** | 5 | 17.8571 | 6 | 7.6923 | 2.5813(0.7093-10.6349) | 0.1544 |
| **DPB1*05:01** | 2 | 7.1429 | 3 | 3.8462 | 1.9098(0.226-12.9377) | 0.6059 |
| **DPB1*06:01** | 2 | 7.1429 | 3 | 3.8462 | 1.9098(0.226-12.9377) | 0.6059 |
| **DPB1*09:01** | 0 | 0.0000 | 1 | 1.2821 | 0(0-52.9286) | 1.0000 |
| **DPB1*10:01** | 0 | 0.0000 | 3 | 3.8462 | 0(0-4.8004) | 0.5642 |

| | | | | | | |
|---|---|---|---|---|---|---|
| **DPB1*11:01** | 1 | 3.5714 | 4 | 5.1282 | 0.6874(0.0273-5.5561) | 1.0000 |
| **DPB1*14:01** | 0 | 0.0000 | 1 | 1.2821 | 0(0-52.9286) | 1.0000 |
| **DPB1*15:01** | 0 | 0.0000 | 1 | 1.2821 | 0(0-52.9286) | 1.0000 |
| **DPB1*16:01** | 0 | 0.0000 | 1 | 1.2821 | 0(0-52.9286) | 1.0000 |
| **DPB1*17:01** | 0 | 0.0000 | 1 | 1.2821 | 0(0-52.9286) | 1.0000 |
| **DPB1*19:01** | 0 | 0.0000 | 2 | 2.5641 | 0(0-9.7321) | 1.0000 |
| **DQA1*01:01** | 4 | 12.5000 | 13 | 16.2500 | 0.7382(0.2053-2.5962) | 0.7742 |
| **DQA1*01:02** | 6 | 18.7500 | 13 | 16.2500 | 1.1874(0.3806-3.5601) | 0.7836 |
| **DQA1*01:03** | 2 | 6.2500 | 10 | 12.5000 | 0.4694(0.0699-2.3032) | 0.5036 |
| **DQA1*02:01** | 5 | 15.6250 | 16 | 20.0000 | 0.7427(0.2384-2.1879) | 0.7896 |
| **DQA1*03:01** | 7 | 21.8750 | 16 | 20.0000 | 1.1188(0.359-3.314) | 1.0000 |
| **DQA1*04:01** | 2 | 6.2500 | 1 | 1.2500 | 5.1757(0.3925-154.52) | 0.1959 |
| **DQA1*05:01** | 6 | 18.7500 | 11 | 13.7500 | 1.4425(0.448-4.7541) | 0.5634 |
| **DQB1*02:01** | 4 | 12.5000 | 7 | 8.5366 | 1.5245(0.3887-5.5554) | 0.7253 |
| **DQB1*02:02** | 4 | 12.5000 | 9 | 10.9756 | 1.1572(0.3075-4.276) | 1.0000 |
| **DQB1*03:01** | 3 | 9.3750 | 12 | 14.6341 | 0.6059(0.1367-2.2411) | 0.5517 |
| **DQB1*03:02** | 5 | 15.6250 | 12 | 14.6341 | 1.0795(0.3363-3.5993) | 1.0000 |
| **DQB1*03:03** | 2 | 6.2500 | 4 | 4.8780 | 1.2969(0.1662-7.3473) | 1.0000 |
| **DQB1*04:02** | 2 | 6.2500 | 1 | 1.2195 | 5.3062(0.4025-158.3838) | 0.1898 |
| **DQB1*05:01** | 2 | 6.2500 | 11 | 13.4146 | 0.433(0.0651-2.0132) | 0.3463 |
| **DQB1*05:02** | 1 | 3.1250 | 1 | 1.2195 | 2.5876(0.0658-101.7662) | 0.4844 |
| **DQB1*05:03** | 2 | 6.2500 | 4 | 4.8780 | 1.2969(0.1662-7.3473) | 1.0000 |
| **DQB1*05:04** | 1 | 3.1250 | 0 | 0.0000 | Inf(0.1349-Inf) | 0.2807 |
| **DQB1*06:01** | 1 | 3.1250 | 1 | 1.2195 | 2.5876(0.0658-101.7662) | 0.4844 |
| **DQB1*06:02** | 2 | 6.2500 | 9 | 10.9756 | 0.5433(0.0799-2.7463) | 0.5128 |
| **DQB1*06:03** | 2 | 6.2500 | 8 | 9.7561 | 0.619(0.0898-2.8862) | 0.7229 |
| **DQB1*06:04** | 0 | 0.0000 | 2 | 2.4390 | 0(0-8.9399) | 1.0000 |
| **DQB1*06:09** | 1 | 3.1250 | 1 | 1.2195 | 2.5876(0.0658-101.7662) | 0.4844 |
| **DRB1*01:01** | 2 | 6.6667 | 8 | 11.1111 | 0.5743(0.0829-2.6834) | 0.7192 |
| **DRB1*03:01** | 4 | 13.3333 | 5 | 6.9444 | 2.0454(0.4832-8.2186) | 0.4433 |
| **DRB1*04:01** | 4 | 13.3333 | 8 | 11.1111 | 1.2282(0.3183-4.823) | 0.7442 |
| **DRB1*04:02** | 1 | 3.3333 | 0 | 0.0000 | Inf(0.1263-Inf) | 0.2941 |
| **DRB1*04:04** | 1 | 3.3333 | 4 | 5.5556 | 0.589(0.0234-4.753) | 1.0000 |
| **DRB1*07:01** | 4 | 13.3333 | 14 | 19.4444 | 0.64(0.1781-2.2209) | 0.5757 |
| **DRB1*08:01** | 2 | 6.6667 | 1 | 1.3889 | 4.9787(0.3766-148.9925) | 0.2061 |
| **DRB1*09:01** | 1 | 3.3333 | 1 | 1.3889 | 2.424(0.0615-95.5137) | 1.0000 |
| **DRB1*10:01** | 1 | 3.3333 | 2 | 2.7778 | 1.2046(0.0403-15.923) | 1.0000 |
| **DRB1*11:01** | 1 | 3.3333 | 5 | 6.9444 | 0.465(0.0192-3.7846) | 0.6677 |
| **DRB1*11:02** | 0 | 0.0000 | 1 | 1.3889 | 0(0-45.6) | 1.0000 |

| | | | | | | |
|---|---|---|---|---|---|---|
| **DRB1*11:04** | 2 | 6.6667 | 1 | 1.3889 | 4.9787(0.3766-148.9925) | 0.2061 |
| **DRB1*12:01** | 0 | 0.0000 | 1 | 1.3889 | 0(0-45.6) | 1.0000 |
| **DRB1*13:01** | 2 | 6.6667 | 7 | 9.7222 | 0.6657(0.0943-3.4203) | 0.7231 |
| **DRB1*13:02** | 1 | 3.3333 | 3 | 4.1667 | 0.7948(0.0297-7.5246) | 1.0000 |
| **DRB1*14:01** | 0 | 0.0000 | 2 | 2.7778 | 0(0-8.3711) | 0.5807 |
| **DRB1*15:01** | 3 | 10.0000 | 7 | 9.7222 | 1.0314(0.2153-4.3008) | 1.0000 |
| **DRB1*15:02** | 0 | 0.0000 | 1 | 1.3889 | 0(0-45.6) | 1.0000 |
| **DRB1*15:03** | 0 | 0.0000 | 1 | 1.3889 | 0(0-45.6) | 1.0000 |
| **DRB1*16:01** | 1 | 3.3333 | 0 | 0.0000 | Inf(0.1263-Inf) | 0.2941 |
